# Increased PCR screening capacity using a multi-replicate pooling scheme

**DOI:** 10.1101/2020.04.16.20067603

**Authors:** A. Viehweger, F. Kühnl, C. Brandt, B. König, A. C. Rodloff

**Affiliations:** Institute for Medical Microbiology and Epidemiology of Infectious Diseases, University Hospital Leipzig; Interdisciplinary Center of Bioinformatics, University Leipzig; Institute for Infectious Diseases and Infection Control, Jena University Hospital; nanozoo GmbH, Leipzig

## Abstract

Effective public health response to viral outbreaks such as SARS-CoV-2 require reliable information about the spread of the infecting agent. Often real-time PCR screening of large populations is a feasible method to generate this information. Since test capacities are usually limited, pooling of test specimens is often necessary to increase screening capacity, provided that the test sensitivity is not significantly compromised. However, when a traditional pool is tested positive, all samples in the pool need individual retesting, which becomes ineffective at a higher proportion of positive samples. Here, we report a new pooling protocol that mitigates this problem by replicating samples across multiple pools. The resulting pool set allows the sample status to be resolved more often than with traditional pooling. At 2% prevalence and 20 samples per pool, our protocol increases screening capacity by factors of 5 and 2 compared to individual testing and traditional pooling, respectively. The corresponding software to layout and resolve samples is freely available under a BSD license (https://github.com/phiweger/clonepool).

## Main text

Pooling of test specimen has long been used to screen larger collections of samples, when most of them are expected to test negative.^1^ In outbreak situations such as the current SARS-CoV-2 pandemic, this “pooling” allows screening of large populations to guide public health response. However, in most labora- tories, the screening capacity is limited by the number of PCR reactions that can be performed in a given time. It is, therefore, desirable to maximize the number of samples that can be tested per reaction.

Various approaches have been proposed to do so in the context of SARS-CoV-2 RT-PCR testing.^2,3^ One problem with the traditional pooling approach, where several samples are collected and tested collectively, is that the number of positive pools that require individual retesting increases rapidly with the number of positive samples in the overall population. A high prevalence of the target renders traditional pooling ineffective. To mitigate this, we propose to test specimens in replicates and distribute them across multiple pools. The resulting “pool address” can then be used to resolve samples in one pool, given the information from other pools that contain a replicate. While some previous studies have taken a similar approach implicitly,^2^ it has neither been investigated systematically for more replicates than two, nor is there any software that would generate and resolve the corresponding pooling layout for laboratory use.

We therefore introduce “clonepool”, a pooling framework to maximize the effective number of samples *s*_*e*_ per PCR reaction. “Effective” refers to the fact that samples in positive pools, whose status cannot be resolved in the pooled run, are assumed to be retested individually. The maximum number of samples for a given pool size *p*, number of pools *n* and number of replicates *r* is calculated as 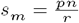. The effective number of samples can then be calculated from the number of unresolved samples *s* as 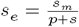.

The clonepool algorithm first distributes all sample replicates randomly across the available pools, with the limitation that a sample’s replicates do not co-occur in the same pool. After the pools have been tested, the algorithm attempts to resolve the samples’ status in two phases: In a first phase, all samples that have at least one replicate in a negative pool are marked negative. In the second phase, samples that (a) only occur in positive pools, and (b) have at least one replicate in a pool in which all other samples are known to be negative, are marked positive (red, orange). All other samples cannot be resolved, and need to be retested individually. The longer the set of pools a sample is distributed across, i.e., the larger its pool set, the more samples can be resolved given a particular prevalence. Of course, a higher number of replicates comes at the price of a reduced number of samples which can be processed at a fixed number of wells. Fortunately, our results provide an efficient means to find the optimal trade-off for any given set of parameters.

We tested the proposed clonepool algorithm using simulated data. We assumed no pipetting errors, which can be achieved, e.g., through the use of a pipetting robot. We also assume that 94 pools are available, which corresponds to a 96-well plate with two wells reserved for a positive and a negative control. Furthermore, we assume that there are no false positive or false negative PCR reactions.

Two parameters determine which pooling scheme is most effective (Fig. 2). If both the prevalence and the number of samples per pool are low, traditional pooling without replicates yields the highest number of samples per reaction. However, as the prevalence increases or more and more samples are pooled, the number of positive pools increases, causing a large number of retested samples and thus reducing the overall throughput. Using sample replicates will then allow to resolve more samples than in the traditional approach. In our testing experience, we observed a prevalence of about 5%, but this value is subject to variability, e.g., depending on a population’s pre-test probability.

**Figure 1:**
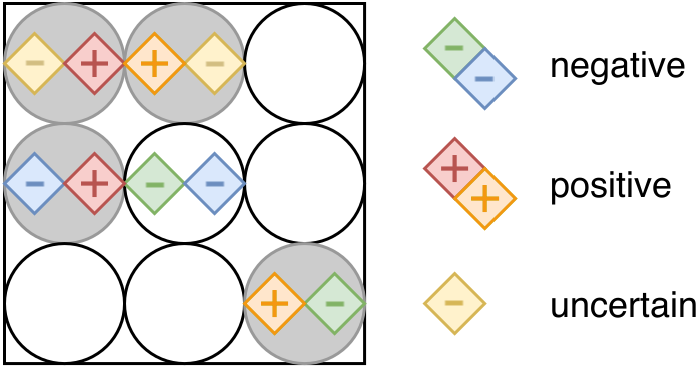
Illustration of the clonepool algorithm. Circles denote the wells, each containing a pool of samples (small squares). A distinct color marks all replicates of a single sample. Positive samples are flagged with “+”, negative ones remain empty. Positive pools are shaded in grey, negative ones in white. In a first phase, all samples that have at least one replicate in a negative pool are identified as negative (blue, green). In the second phase, samples that only occur in positive pools and where at least one replicate is in a pool where all other samples are negative, are recognized as positive (red, orange). All other samples cannot be resolved and have to be retested individually (yellow).

**Figure 2:**
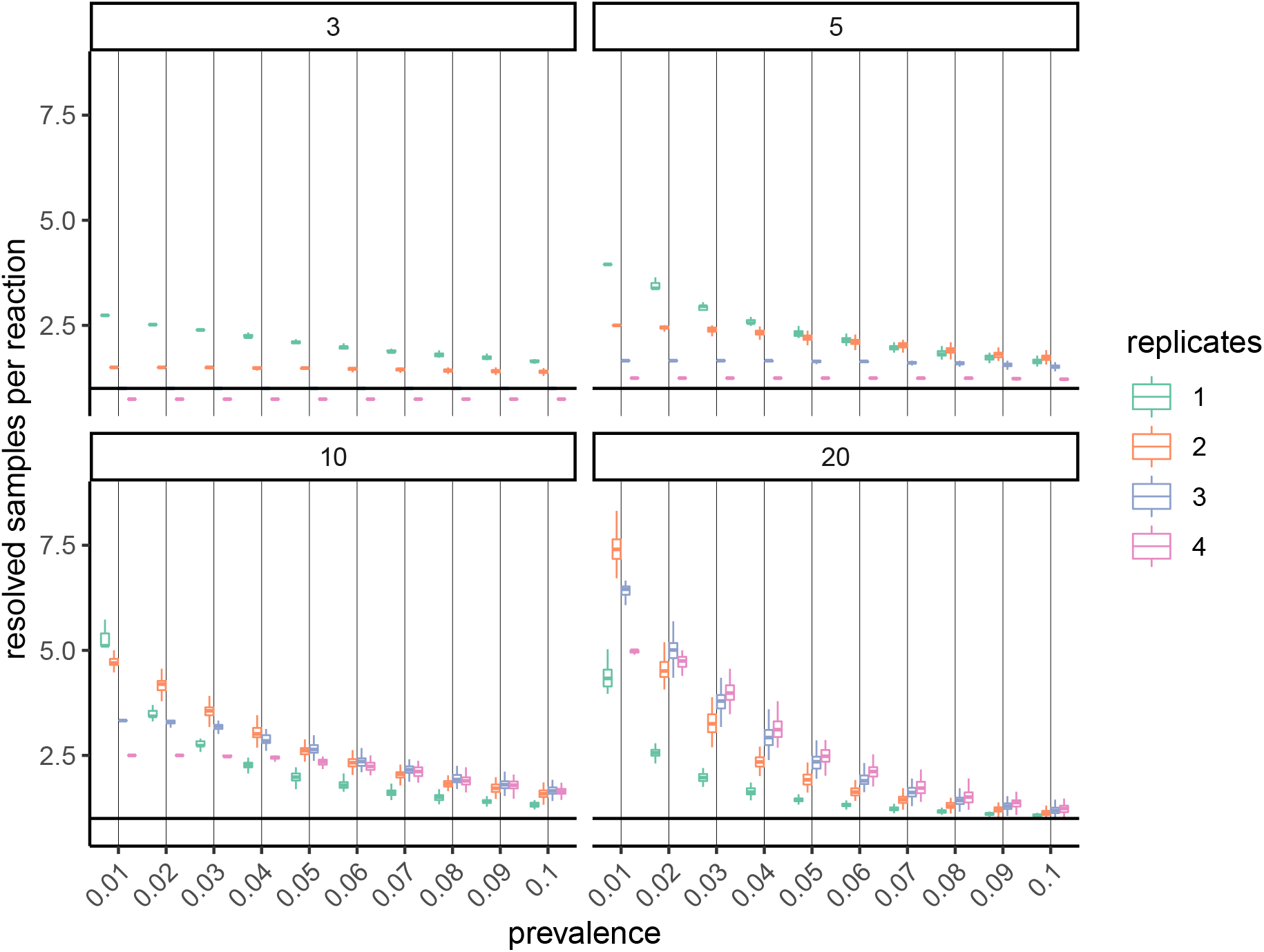
Simulation results for different percentages of positive samples (x-axis), replicates (colors), and pool sizes (panels). The target metric is the effective number of samples per PCR reaction, which includes the individual retesting of samples that cannot be resolved in the first pooling run.

The number of samples that can be pooled without affecting the PCR sensitivity is limited by the PCR cycle threshold (Ct) for the target, i.e., the cycle at which amplification becomes detectable over background noise (typically ten times the standard deviation, SD). Usually, Ct values above 35 are treated as unspecific amplification. SARS-CoV-2 amplifies at low Ct values due to high viral titers (Ct 18-25 depending on the material and number of days post-infection).^4,5^ A 20-fold dilution, i.e., pooling 20 samples, would cause the Ct value to increase by about 4.3 cycles (2^*x*^ = *d*, where d is the dilution and × the shift in Ct), which still lies comfortably above the detection limit.

At a prevalence of 5% SARS-CoV-2 positive samples, and for ten samples per pool and two replicates per sample, we simulate that 2.61 times the number of samples can be processed compared to testing samples individually (SD 0.13). This result is in line with previous estimates using a slightly different version of the 2-replicate scheme.^2^ Using two replicates increases the effective number of samples per reaction by 31% compared to pooling without replicates. At 2% prevalence and 20 samples per pool – a scenario more akin to screening large populations – 5.01 times the number of samples can be screened compared to individual testing (SD 0.28), and the increase over traditional pooling is 193%. These presented values correspond to *in silico* simulations, and require further validation in the laboratory.

In conclusion, our pooling protocol based on sample replicates can substantially increase the number of samples per PCR reaction when screening large populations during pathogen outbreaks, such as SARS- CoV-2. The protocol can be tuned to local laboratory conditions such as pool size and proportion of positive samples. Also, subsequent testing of positive pools could be made more efficient by splitting pools iteratively.^6^

The clonepool software supports the protocol’s implementation for routine use (https://github.com/phiweger/clonepool).

## Data Availability

The corresponding software to layout and resolve samples is freely available under a BSD license (https://github.com/phiweger/clonepool).

